# Serum interferon-gamma-induced protein 10 levels can help predict sarcopenia development in patients with primary hepatocellular carcinoma: A retrospective cohort study

**DOI:** 10.1101/2024.08.21.24312385

**Authors:** Hitomi Takada, Leona Osawa, Yasuyuki Komiyama, Masaru Muraoka, Yuichiro Suzuki, Mitsuaki Sato, Shoji Kobayashi, Takashi Yoshida, Shinichi Takano, Shinya Maekawa, Nobuyuki Enomoto

## Abstract

**Background:** Sarcopenia is a prognostic factor in patients with hepatocellular carcinoma (HCC). However, the mechanism underlying sarcopenia development in these patients remains unclear. The chemokine interferon-gamma-induced protein 10/C-X-C motif chemokine ligand 10 (IP-10) has been found to be associated with muscle regeneration or destruction. Thus, we aimed to clarify the role of serum IP-10 levels in predicting sarcopenia development in patients with HCC.

**Methods:** This retrospective study enrolled 120 patients with primary HCC whose serum IP-10 levels were measured both at baseline and 1 year after the confirmed diagnosis of HCC. Patients who had sarcopenia at baseline computed tomography imaging were assigned to the Sarco-base group, whereas those in whom sarcopenia was found for the first time during the 3-year follow-up were assigned to the Sarco-develop group. Those who never met the criteria during the follow-up period were assigned to the Non-Sarco group.

**Results:** The baseline IP-10 levels were significantly lower in the Sarco-base group compared to the rest (88 vs. 110 pg/ml, p = 0.016). Conversely IP-10 levels and IP-10 ratio at 1 year after the confirmed diagnosis of HCC were lower in the Non-Sarco group compared to the rest (25 vs. 62 pg/ml, p < 0.001, 0.91 vs. 1.1, p = 0.044). Further comparisons between the three groups show that the baseline IP-10 levels were higher in the Sarco-develop group than in the Sarco-base (p < 0.001) and Non-Sarco (p = 0.0017) groups.

**Conclusions:** Patients with sarcopenia at baseline more frequently presented with low IP-10 levels than those without. Contrarily, the group without sarcopenia at baseline and with high baseline IP-10 levels and IP-10 ratios at 1 year were more likely to develop sarcopenia within 3 years. Monitoring of IP-10 levels may enable the identification of groups prone to develop sarcopenia in patients with HCC.

## Introduction

Sarcopenia is characterized by a low skeletal muscle mass, weakness, and decreased overall physical performance [1]. Based on the evaluation criteria developed by the Japan Society of Hepatology (JSH) for sarcopenia in liver disease cases, patients with chronic liver disease exhibiting low grip strength and muscle mass, as determined by computed tomography (CT) or bioelectrical impedance analysis, are considered to have sarcopenia [2]. Recently, there has been a growing emphasis on evaluating sarcopenia in terms of muscle quality, especially focusing on decreased grip strength or increased intramuscular fat mass [3–6]. Sarcopenia reportedly is a poor prognostic factor in patients with hepatocellular carcinoma (HCC) [7–11]. Thus, sarcopenia has recently gained more attention in those with chronic liver disease.

However, the development of sarcopenia in patients with HCC is often associated with primary sarcopenia due to age and secondary sarcopenia, making it difficult to understand the pathogenesis in some cases. Several theories about sarcopenia development, besides primary sarcopenia, have been proposed; however, the precise mechanism in patients with HCC still requires elucidation. The decreased levels of amino acids and testosterone, hyperammonemia, hypermetabolic state, and suppression of the mammalian target of rapamycin pathway due to lack of mobility and systemic treatment are all thought to contribute to development of sarcopenia [12].

Chemokine interferon-gamma (IFN-γ)-induced protein 10/C-X-C motif chemokine ligand 10 (IP-10), a downstream molecule of IFN-γ, may potentially be involved in the mechanism [13, 14]. IP-10 levels were reported to promote the proliferation and differentiation of satellite cells by signaling through the receptor CXCR3. Intramuscular recombinant IP-10 treatment in aged mice induces the proliferation of satellite cells and provokes the increase in the number of regenerated myofibers [15]. Contrarily, the reduction in muscle atrophy via down-regulation of IP-10 levels has been reported in tumor-bearing mice [16]. The serum IP-10 levels correlated with the clinical degree of muscle involvement in patients with systemic sclerosis, suggesting that high IP-10 levels do not usually indicate increased muscle regeneration [17]. The IP-10 levels may produce an opposite effect on muscle regeneration depending on age, presence of chronic disease, presence of cancer, and so on. Therefore, in the present study, we aimed to examine the association between serum IP-10 levels and sarcopenia development in patients with HCC.

## Methods

### Patients

From a total of 738 patients at our hospital with a confirmed diagnosis of primary HCC from January 2008 to January 2021, 120 patients whose serum IP-10 levels were measured both at baseline and after 1 year, were enrolled in our research. Patients with Barcelona Clinic Liver Cancer (BCLC) stage A who had satisfactory imaging at baseline and were aged ≥20 years were included in the analysis, whereas those without sufficient blood samples for IP-10 assay, those with missing data, those diagnosed with HCC other than BCLC stage A, or those with a shorter follow-up (<3 years) were excluded from the analysis.

HCC was diagnosed based on the outcomes of the pathological evaluation or the non-rim hyperenhancement in the arterial phase of dynamic CT or gadolinium ethoxybenzyl diethylenetriamine penta-acetic acid-contrast-enhanced magnetic resonance imaging (MRI) and non-peripheral washout or threshold increase, where only nodules that exhibited LR-4 and LR-5 using Liver Imaging Reporting and Data System were categorized as HCC [18].

All patients provided written informed consent prior to study participation, and the study was approved by the Human Ethics Review Committee of Yamanashi University Hospital (approval number: 1326), in accordance with the Declaration of Helsinki.

### Diagnosis of sarcopenia

According to the evaluation criteria for sarcopenia in liver disease established by JSH, the manifestation of decreased grip strength and low muscle mass are indicative of sarcopenia. Nonetheless, due to the retrospective nature of our research, adequate grip strength analysis was not possible. Hence, CT values were used for assessing skeletal muscle quality.

The psoas muscle mass index (PMI) at the level of the third lumbar vertebra, as determined by CT imaging, was applied as an indicator of muscle mass volume. CT images obtained during the primary HCC diagnosis served as the baseline data. The cross-sectional areas of the bilateral psoas muscles were evaluated by manual tracing, and PMI was determined by normalizing these areas to a square of a patient’s height in meters. The cut-off value for PMI was set as 6.36 and 3.92 cm^2^/m^2^ for men and women, respectively, as per the JSH criteria [19]. The CT values for the multifidus muscle at the third lumbar vertebral level were indicative of skeletal muscle quality [20–22]. The cut-off value for low CT values was established at 44.4 and 39.3 Hounsfield Unit (HU) for men and women, respectively [3, 4]. The measurement of muscle mass volume and CT values was carried out by two hepatology specialists who were experts in this field.

In our research, sarcopenia was outlined as the presence of both low PMI and CT values. Patients with sarcopenia at baseline CT at the confirmed diagnosis of HCC were assigned to the Sarco-base group, those meeting the criteria for sarcopenia for the first time at the 3-year follow-up were referred to as the Sarco-develop group, and those never meeting the criteria during follow-up were assigned to the Non-Sarco group.

### Evaluation of serum IP-10 levels

Serum samples were gathered from 9 mL of blood obtained at baseline and 1 year after the confirmed diagnosis of HCC. These samples were distributed into aliquots and stored at −80 °C until further analysis. The serum IP-10 levels were measured using 50-μL of the stored serum and an enzyme-linked immunosorbent assay kit, as per the manufacturer’s instructions. The levels were determined using standard calibration curves and expressed in pg/mL. The IP-10 ratio was defined as the ratio of IP-10 levels obtained at baseline and 1 year after the confirmed diagnosis of HCC.

### Statistical analysis

All experimental data were expressed as medians (ranges). Between-group comparisons were conducted using the Mann–Whitney U-, Kruskal–Wallis, and Friedman tests along with nonparametric analysis of variance. If the one-way analysis of variance yielded significant results, differences between individual groups were analyzed with Fisher’s exact test. A *p* < 0.05 was regarded as statistically significant. All the analyses were done with EZR (Saitama Medical Center, Jichi Medical University, Saitama, Japan), a graphical user interface for R (The R Foundation for Statistical Computing, Vienna, Austria). Specifically, EZR is a modified version of the R commander developed to employ statistical functions commonly applied in biostatistics [23].

## Results

### Patient characteristics

The median age of the patients (n = 120) was 71 years (range: 51—87 years), and 73 patients were male. The Child–Pugh scores were 5, 6, and 7 in 86, 22, and 12 cases, respectively. Regarding the Tumor-Node-Metastasis (TNM) staging, 52 and 68 patients were categorized as having Stage 1 and 2, respectively. Forty patients did not experience sarcopenia (Non-Sarco group), 40 had sarcopenia during the 3-year follow-up (Sarco-develop group), and 40 patients were assigned to the Sarco-base group. A summary of the features of these patients is shown in Table 1.

**Table 1.** Baseline characteristics of 120 BCLC stage A patients.

|  | All (n=120) |
| --- | --- |
| Age, years old | 71 (51-87) |
| Male, n | 75 (63%) |
| Body mass index | 23 (15-40) |
| Diabetes mellitus, n | 44 (37%) |
| Habitual drinker, n | 12 (10%) |
| Etiology (viral/nonviral), n | 93/27 (78/23%) |
| Child-Pugh grade (A/B), n | 109/11 (91/9%) |
| Albumin-bilirubin grade (1/2a/2b), n | 59/34/27 (49/28/23%) |
| Branched-chain amino acids, $\mu\text{mol/ml}$ | 463 (276-973) |
| Branched-chain amino acids/tyrosine ratio | 5.3 (2.4-11) |
| Platelet, $\times 10^3/\mu\text{l}$ | 115 (53-258) |
| Alpha-fetoprotein, ng/ml | 4.8 (1.4-6800) |
| Des-γ-carboxy prothrombin, mAU/ml | 21 (8.0-7397) |
| Tumor size, maximum, mm | 19 (8.0-55) |
| The number of intrahepatic tumors, n | 1 (1-3) |
| Up to seven beyond at baseline, n | 9 (7.5%) |
| Therapy for primary HCC (resection/ percutaneous puncture treatment/ transarterial chemoembolization), n | 29/51/40 (24/43/33%) |
| IP-10/CXCL10 at baseline, pg/ml | 100 (36-456) |

### Characteristics of IP-10 levels in BCLC stage A patients

The IP-10 levels only showed a very weak correlation with ALBI score, Child-Pugh score, Branched-chain amino acids (BCAA), Branched-chain amino acids/tyrosine ratio (BTR), platelet counts, alpha-fetoprotein (AFP), and tumor size, with no alternative indicators (Table 2).

**Table 2.**
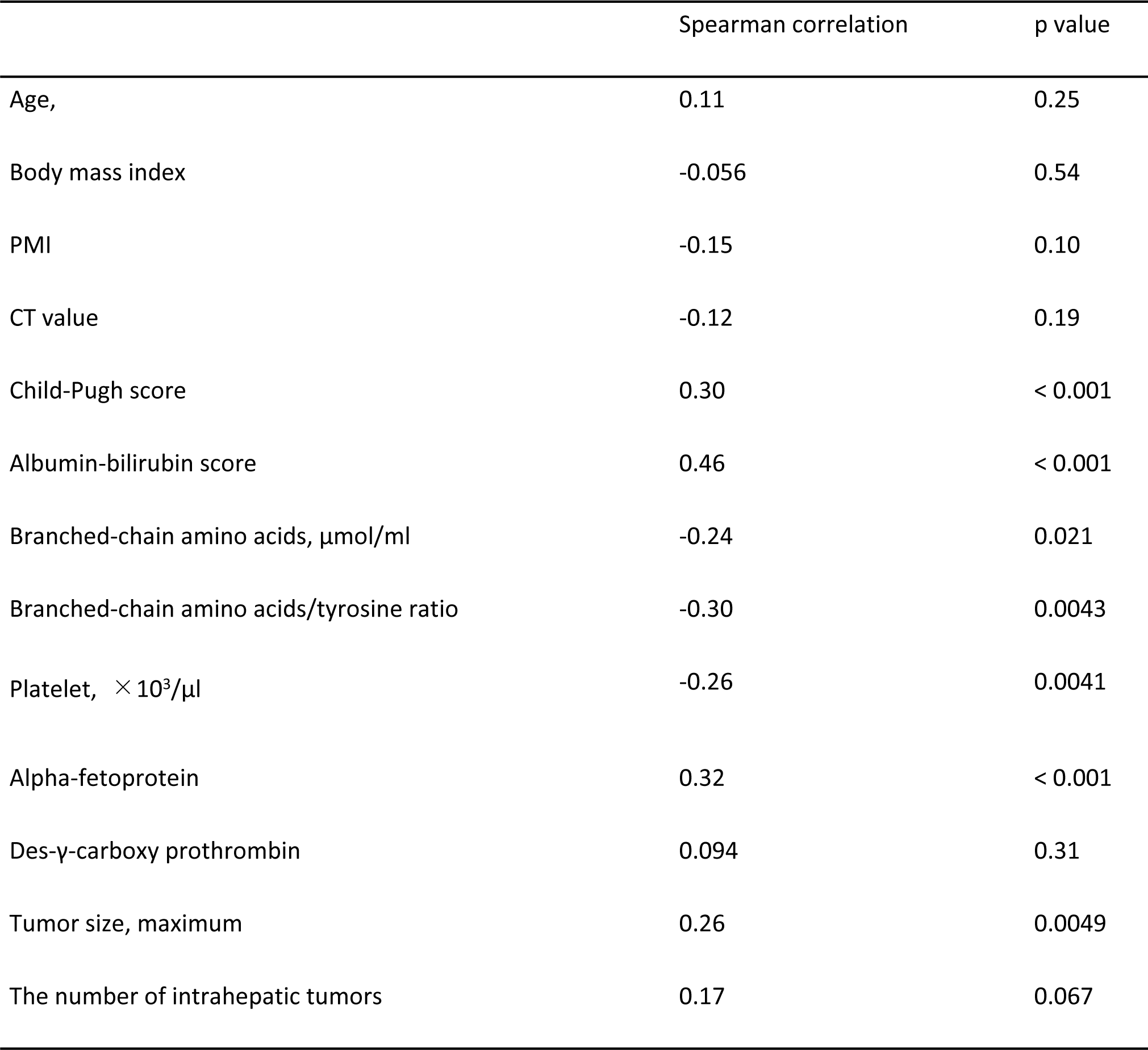
Factors associated with IP-10 levels in BCLC stage A patients.

### Association between IP-10 levels and sarcopenia development

The baseline IP-10 levels were significantly lower in the Sarco-base group compared to the rest (88 vs. 110 pg/ml, p = 0.016) (Figure 1a). No association was found between the presence of sarcopenia at baseline and IP-10 ratio (0.98 vs. 1.0, p =0 .81) (Figure.1b). The comparative analysis of the characteristics of patients stratified according to the diagnosis of sarcopenia at baseline are reflected in Table 3. Contrarily, IP-10 levels at 1 year after the confirmed diagnosis of HCC were lower in the Non-Sarco group compared to the rest (25 vs. 62 pg/ml, p < 0.001) (Figure.1c). The IP-10 ratio after 1 year was also lower in the Non-Sarco group compared to the rest (0.91 vs. 1.1, p = 0.044) (Figure.1d).

**Figure 1.**
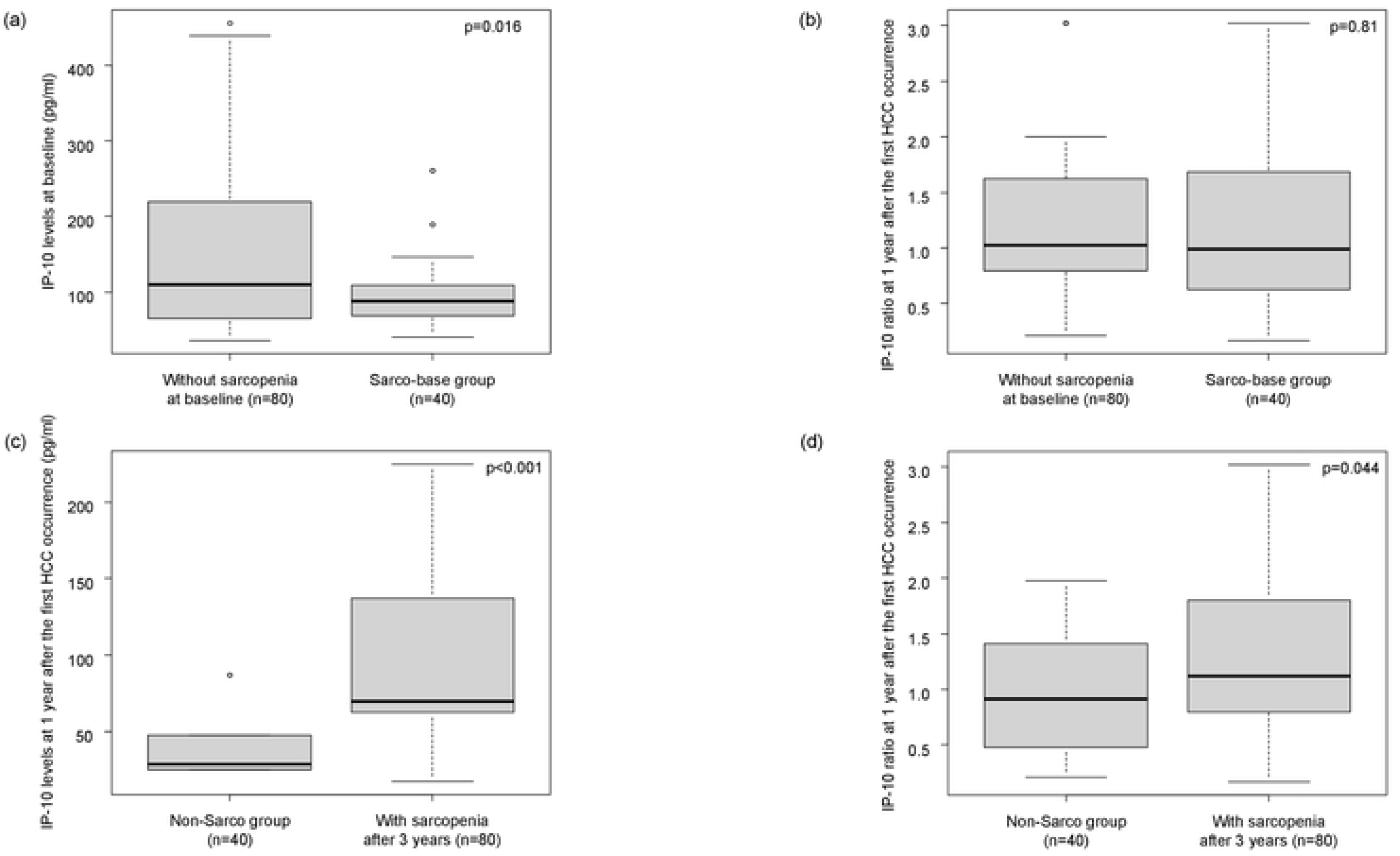
Association between IP-10 levels and sarcopenia development. (a) The association between serum IP-10 levels and the presence of sarcopenia at baseline, (b) The association between IP-10 ratios and the presence of sarcopenia at baseline. (c) The association between serum IP-10 levels at 1 year after the first HCC occurrence and sarcopenia during 3 follow-up years. (d) The association between IP-10 ratios at 1 year after the first HCC occurrence and sarcopenia during 3 follow-up years.

**Table 3.** Characteristics of patients with comparison made by the presence of sarcopenia at baseline.

|  | With sarcopenia at baseline (n=40) | Without sarcopenia at baseline (n=80) | p value |
| --- | --- | --- | --- |
| Age, years old | 74 (56-83) | 71 (51-87) | 0.46 |
| Male, n | 34 (85%) | 38 (48%) | < 0.001 |
| Body mass index | 22 (17-29) | 24 (15-40) | 0.013 |
| Diabetes mellitus, n | 10 (25%) | 37 (46%) | 0.030 |
| Habitual drinker, n | 2 (5.0%) | 7 (8.8%) | 0.72 |
| Etiology (viral/nonviral), n | 34/6 (85/15%) | 59/21 (74/26%) | 0.25 |
| Child-Pugh grade (A/B), n | 36/4 (90/10%) | 60/20 (75/25%) | 0.32 |
| Albumin-bilirubin grade (1/2a/2b), n | 22/10/8 (55/25/20%) | 36/23/21 (45/29/26%) | 0.59 |
| Branched-chain amino acids, $\mu\text{mol/ml}$ | 456 (311-973) | 449 (262-636) | 0.98 |
| Branched-chain amino acids/tyrosine ratio | 5.0 (2.9-11) | 5.3 (2.4-8.4) | 0.59 |
| Platelet ($\times 10^3/\mu\text{l}$) | 129 (57-258) | 107 (53-382) | 0.077 |
| Alpha-fetoprotein, ng/ml | 5.1 (1.4-6800) | 5.9 (1.2-2210) | 0.97 |
| Des-γ-carboxy prothrombin, mAU/ml | 22 (11-7397) | 20 (8.0-18108) | 0.48 |
| Tumor size, maximum, mm | 19 (8.0-60) | 18 (9.0-70) | 0.68 |
| The number of intrahepatic tumors, n | 1 (1-2) | 2 (1-3) | < 0.001 |
| Up to seven beyond at baseline, n | 2 (5.0%) | 11 (14%) | 0.22 |
| Therapy for primary HCC (resection/ percutaneous puncture treatment/ transarterial chemoembolization), n | 6/24/10 (15/60/25%) | 23/27/30 (29/33/38%) | 0.036 |
| IP-10 level at baseline, pg/ml | 88 (40-261) | 110 (2.5-527) | 0.016 |

### Comparison of the association between IP-10 levels and sarcopenia development among the three patient groups

Further comparisons were performed between the Sarco-base, Sarco-develop, and Non-Sarco groups. The baseline IP-10 levels were higher in the Sarco-develop group than in the Sarco-base (p < 0.001) and Non-Sarco (p = 0.0017) groups (Figure 2a). The IP-10 ratios were higher in the Sarco-develop group than in the Non-Sarco group (p = 0.025) (Figure.2b). The outcomes of the comparative analysis of the patient characteristics among the three groups are summarized in Table 4.

**Figure 2.**
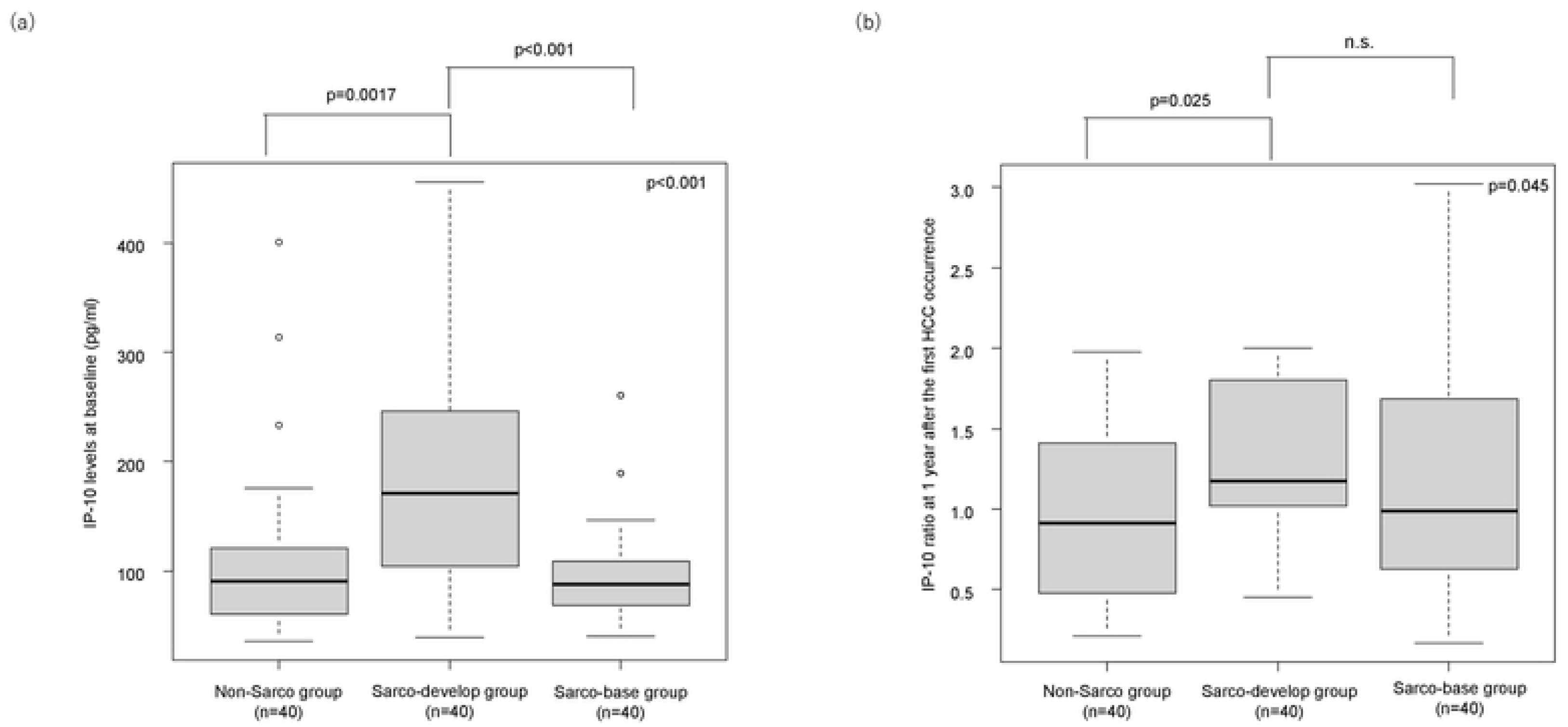
Association between IP-10 levels and sarcopenia development among the three patient groups. (a) Baseline IP-10 levels among three groups. (b) IP-10 ratios among three groups.

**Table 4.**
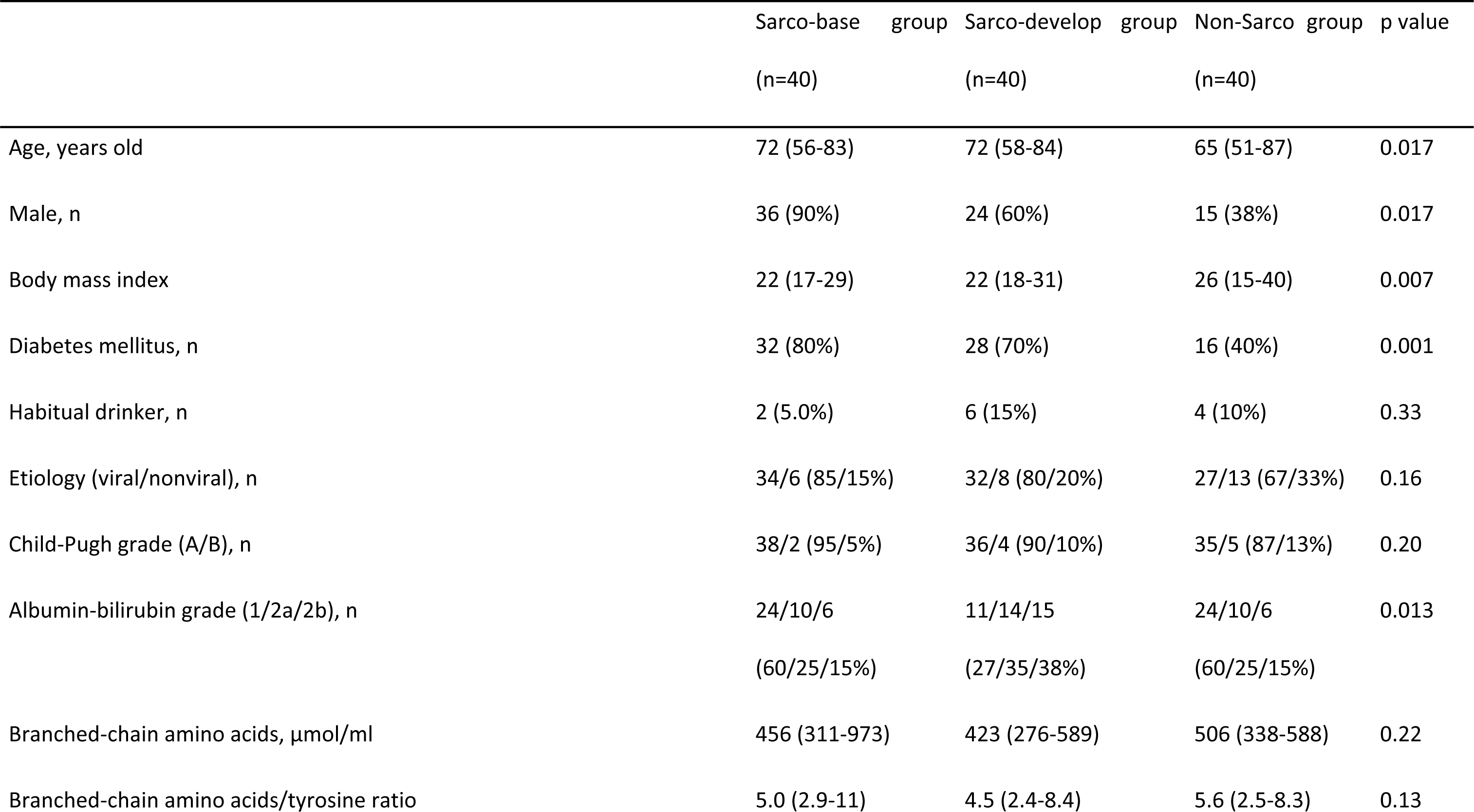
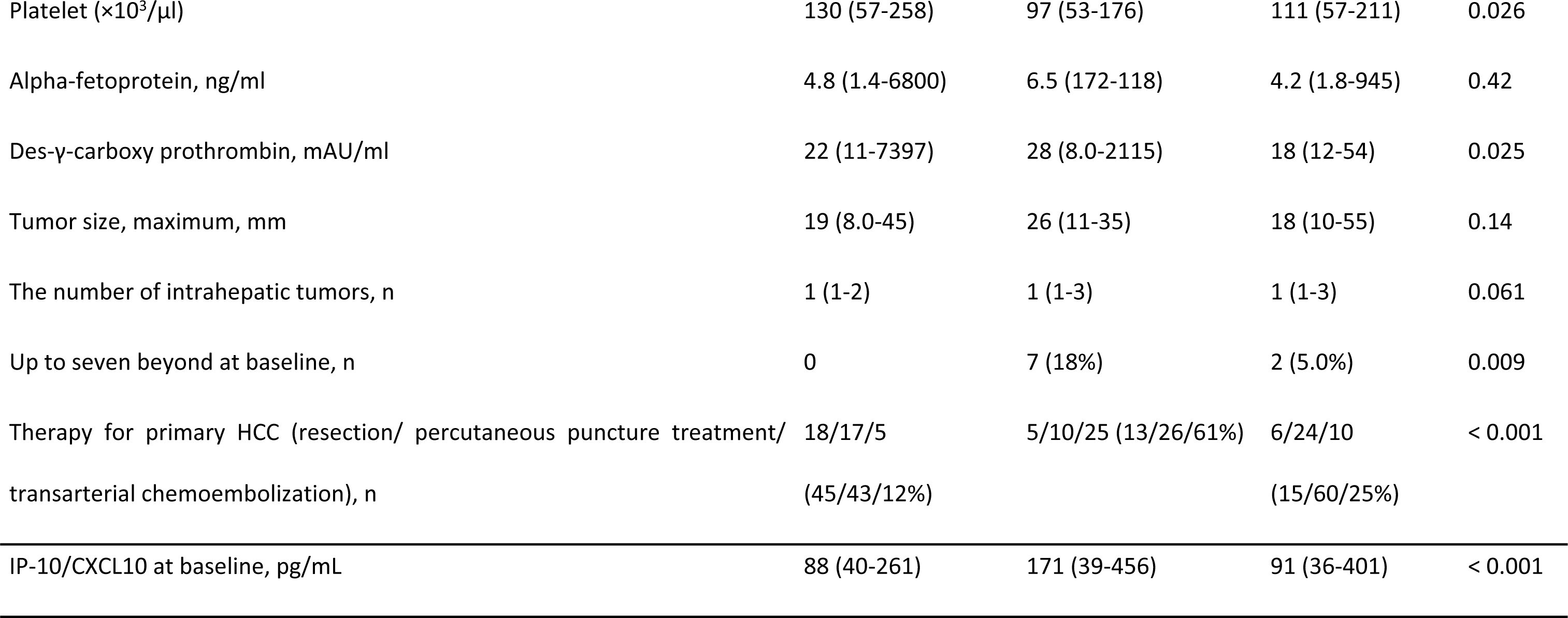
Characteristics of patients with comparison made by three groups.

### Characteristics of patients with high baseline IP-10 levels or ratios

Patients without sarcopenia at baseline but who had high IP-10 levels at baseline or IP-10 ratios after 1 year exhibited several distinguishing characteristics, which were as follows: high ALBI grade, low BCAA, low BTR, low platelet counts, more patients opting for TACE as treatment for primary HCC, and recurrence beyond up to seven criteria during the 3-year follow-up period (Table 5).

**Table 5.** Characteristics of patients presenting high IP-10 levels at baseline or high IP-10 ratios, without sarcopenia at baseline.

|  | The rest of patients (n=22) | Patients with high baseline IP-10 levels or 1-year ratios (n=58) | p value |
| --- | --- | --- | --- |
| Age, years old | 67 (51-75) | 71 (52-87) | 0.58 |
| Male, n | 12 (55%) | 27 (47%) | 0.62 |
| Body mass index | 24 (15-31) | 24 (18-40) | 0.39 |
| Diabetes mellitus, n | 12 (55%) | 22 (41%) | 0.32 |
| Habitual drinker, n | 0 | 10 (17%) | 0.054 |
| Etiology (viral/nonviral), n | 16 (73%) | 43 (74%) | 1.0 |
| Child-Pugh grade (A/B), n | 22/0 (100/0%) | 49/9 (84/16%) | 0.069 |
| Albumin-bilirubin grade (1/2a/2b), n | 16/6/0 (73/27/0%) | 19/18/21 (33/31/36%) | < 0.001 |
| Branched-chain amino acids, $\mu\text{mol/ml}$ | 506 (423-568) | 444 (276-589) | 0.020 |
| Branched-chain amino acids/tyrosine ratio | 5.8 (3.3-6.6) | 4.9 (2.4-8.4) | 0.008 |
| Platelet , $\times 10^3/\mu\text{l}$ | 127 (84-206) | 94 (53-211) | 0.015 |
| Alpha-fetoprotein, ng/ml | 4.2 (1.8-93) | 6.3 (1.7-945) | 0.10 |
| Des-γ-carboxy prothrombin, mAU/ml | 18 (12-49) | 21 (8.0-2115) | 0.41 |
| Tumor size, maximum, mm | 25 (13-55) | 18 (10-48) | 0.060 |
| The number of intrahepatic tumors, n | 1 (1-3) | 1 (1-3) | 0.20 |
| Up to seven beyond at baseline, n | 0 | 9 (16%) | 0.057 |
| Therapy for primary HCC (resection/<br>percutaneous puncture treatment/<br>transarterial chemoembolization), n | 14/8/0 (64/36/0%) | 9/19/30 (15/33/52%) | < 0.001 |
| Recurrence during the 3-year follow-up period, n | 7 (32%) | 29 (50%) | 0.21 |
| Up to seven beyond recurrence during the 3-year follow-up period, n | 0 | 17 (29%) | 0.004 |

## Discussion

In this study, we analyzed the relationship between sarcopenia and serum IP-10 levels in patients with BCLC stage A HCC. Our findings demonstrated that patients with sarcopenia at baseline had lower IP-10 levels than those without. Contrarily, in patients without sarcopenia at baseline, sarcopenia was more likely to develop in those with higher baseline IP-10 levels and 1-year IP-10 ratios. This indicates that high IP-10 levels have an opposite effect on muscle regeneration, depending on the timing after the confirmed diagnosis of HCC. To the best of our knowledge, our results clarified for the first time the relationship between sarcopenia and IP-10 levels in patients with primary HCC, and our data may aid in the identification of those patients at a high-risk of developing sarcopenia and contribute to the development of therapeutic intervention methods.

The term sarcopenia was first introduced in 1989, and since then, numerous studies have analyzed the correlation between sarcopenia and the prognosis of patients with HCC [8, 9, 11, 24]. Nonetheless, the molecular mechanisms serving as a basis of the development of sarcopenia in patients with HCC are complex and have not been completely understood. Particularly, the relationship between IP-10 levels and sarcopenia development in patients with HCC has not been thoroughly investigated.

Previous papers have reported higher peripheral IP-10 concentrations in the elderly than in the younger patients [25]. A decrease in muscle atrophy and tumor shrinkage via down-regulation of the IP-10 levels have been demonstrated in tumor-bearing mice [16]. IFN-γ may facilitate muscle damage by inhibiting M2 macrophage activation and muscle cell proliferation in a mouse model of muscular dystrophy [26]. Serum IP-10 levels were also shown to be correlated with the severity of muscle involvement in patients with systemic sclerosis [17]. Conversely, gene expression patterns associated with the responses to IFN-γ was considerably downregulated during muscle regeneration in aged mice, which is thought to promote satellite cell dysfunctions in aged skeletal muscles [15]. Intramuscular recombinant IP-10 treatment in aged mice provoked the proliferation of satellite cells and caused an increase in regenerated myofibers [15]. IP-10 activated myogenic differentiation in vitro, indicative of its possible direct influence on muscle regeneration [27]. Thus, it has been proposed that IP-10 levels may have an opposing effect on muscle regeneration, and that the effects may also vary according to the presence of malignancies and chronic diseases.

Indeed, the interpretation of the relationship between IP-10 levels and sarcopenia in patients with HCC poses substantial difficulties. Previous studies have indicated an association between the IP-10 levels and cancer stage, with higher levels detected in patients with advanced HCC. The overexpression of IP-10 has been linked to serum AFP levels, tumor size and number along with TNM stage [28]. Patients with higher IP-10 expression levels also had considerably lower overall and disease-free survival rates [29]. The downregulation of IP-10 can suppress metastasis and tumor invasion in HCC patients [28]. Furthermore, the IP-10 levels reportedly are associated with liver function, disease progression, and cirrhosis. Understanding the IP-10 dynamics in patients with chronic hepatitis C may be useful for predicting liver function after direct-acting antiviral therapy [30]. In patients with chronic hepatitis B, the IP-10 levels increased in the group with declining liver function [31]. The IP-10 levels in patients with non-alcoholic steatohepatitis were higher than in those in the control group and patients with non-alcoholic fatty liver [32]. Herein, the IP-10 levels were also found to be correlated with ALBI and Child–Pugh scores, and high IP-10 levels at baseline or IP-10 ratios were associated with high ALBI grade, low BCAA, low BTR, low platelet count, more patients opting for TACE as treatment for primary HCC, and recurrence beyond up to seven criteria during the follow-up period. These results indicate that high IP-10 levels may be associated with advanced cirrhosis or HCC. However, as the analysis by etiology was insufficient in this study, future analysis involving a larger number of cases is needed.

The present study delved into the baseline IP-10 levels and their changes as a potential molecular mechanism underlying sarcopenia development. The low baseline IP-10 levels may indicate that the baseline sarcopenia was due to the decreased levels of IP-10, a factor facilitating muscle differentiation, as described above, or it may reflect muscular atrophy due to sarcopenia, causing an inability to secrete IP-10, presumably a myokine, which is produced by skeletal muscles [14, 15, 27]. Contrarily, the intramuscular CD4 T-cells are increased in both young and aged mice during a viral infection, whereas the number of intramuscular CD8 T-cells increased only in aged muscle [33]. Similar mechanisms may be at work in patients with advanced cirrhosis or poorly controlled HCC. Moreover, it has been proposed that an increasing percentage of CXCR3 variants over time might be an essential component of endometriosis-related carcinogenesis in patients with ovarian cancer [34]. Our findings suggest that higher IP-10 ratios may be associated with sarcopenia development via the dysregulation of T-cell transfer to the muscle, abnormalities in T-cell differentiation, and receptor abnormalities in patients with BCLC stage A.

The present study has some limitations. First, it had a single-center retrospective design and lacked grip strength measurements. Thus, integrating grip strength evaluations, required by the currently widely used guidelines for sarcopenia diagnosis in Japan, is a challenge in future. Second, patient selection bias may have been present. Third, the relationship between etiology and IP-10 levels has not been fully analyzed. The study should be re-evaluated in the future with a larger number of cases.

The present study is the first to demonstrate that the IP-10 levels at baseline or IP-10 ratios may be associated with the development of sarcopenia in patients with BCLC stage A HCC. By evaluating the IP-10 levels, this study not only provides data for identifying high-risk groups for sarcopenia development but also improves our knowledge on the underlying mechanisms involved in sarcopenia development in patients with primary HCC.

## Conclusion

Patients with sarcopenia at baseline were more likely to present with low IP-10 levels than those without. Conversely, those not presenting with sarcopenia at the first occurrence of HCC and subsequently developed sarcopenia within 3 years had higher baseline IP-10 levels and IP-10 ratios at 1 year. This indicates that the IP-10 levels may have different effects on the muscles depending on the timing after the confirmed diagnosis of HCC. Furthermore, monitoring the IP-10 levels may be an effective tool for defining high-risk groups in patients with primary HCC, with a predisposition to develop sarcopenia, which is a significant prognostic factor for patients with primary HCC.

## Data availability

Raw data are available from the corresponding author (H.T.) on reasonable request.

## Funding

This work was not supported by any grant or funding source.

## Ethical statement

The study was conducted in accordance with the ethical guidelines of the 1975 Declaration of Helsinki (6th revision, 2008). The study was approved by the Institutional Review Board (or Ethics Committee) of University of Yamanashi (protocol code 1326). Written informed consent was obtained from all participants.

## Conflicts of Interest

The authors declare no conflict of interest.

## Acknowledgements

We thank Ms. Takako Ohmori and Ms. Tomoko Nakajima for their valuable technical assistance.

## Author contribution

Hitomi Takada: Conceptualization, Data curation, Formal analysis, Methodology, Validation, Visualization, Writing – original draft. Leona Osawa, Yasuyuki Komiyama, Masaru Muraoka, Yuichiro Suzuki, Mitsuaki Sato, Shoji Kobayashi, Takashi Yoshida, and Shinichi Takano: Data curation. Shinya Maekawa and Nobuyuki Enomoto: Supervision.

## Notes

### Competing Interest Statement

The authors have declared no competing interest.

### Funding Statement

The author(s) received no specific funding for this work.

### Author Declarations

the Human Ethics Review Committee of Yamanashi University Hospital (approval number: 1326)

